# The recent rapid expansion of multidrug resistant strains of *Mycobacterium tuberculosis* Ural lineage 4.2 in the Republic of Moldova

**DOI:** 10.1101/2023.11.10.23298377

**Authors:** Melanie H. Chitwood, Caroline Colijn, Chongguang Yang, Valeriu Crudu, Nelly Ciobanu, Alexandru Codreanu, Jaehee Kim, Isabel Rancu, Kyu Rhee, Ted Cohen, Benjamin Sobkowiak

## Abstract

The projected trajectory of multidrug resistant tuberculosis (MDR-TB) epidemics depends on the reproductive fitness of circulating strains of MDR *M. tuberculosis (Mtb)*. Previous efforts to characterize the fitness of MDR *Mtb* have found that *Mtb* strains of the Beijing sublineage (Lineage 2.2.1) may be more prone to develop resistance and retain fitness in the presence of resistance-conferring mutations than other lineages. Using *Mtb* genome sequences from all culture-positive cases collected over two years in Moldova, we estimate the fitness of Ural (Lineage 4.2) and Beijing strains, the two lineages in which MDR is concentrated in the country. We estimate that the fitness of MDR Ural strains substantially exceeds that of other susceptible and MDR strains, and we identify several mutations specific to these MDR Ural strains. Our findings suggest that MDR Ural *Mtb* has been transmitting efficiently in Moldova and poses a substantial risk of spreading further in the region.

## Introduction

Multidrug resistant tuberculosis (MDR-TB) is an important driver of TB-related morbidity and mortality and a major threat to TB control in a number of settings. In several countries of the former Soviet Union upwards of 20% of new TB cases have MDR-TB^1^, with the vast majority of new cases of MDR-TB resulting from direct transmission rather than through resistance acquired during treatment^2^. Determining the epidemiological and genomic factors that influence the spread of these strains is essential to interrupt the transmission of MDR-TB.

The ability for *Mycobacterium tuberculosis* (*Mtb*) to survive, reproduce, and transmit can vary greatly among drug-resistant strains^3^. Drug resistance in *Mtb* is conferred through chromosomal mutation and is often associated with reductions in bacterial fitness in the absence of the selective pressure of antibiotic treatment. However, the degree and the durability of these fitness costs, as well as the rate of acquisition of resistance-conferring mutations^4^, appear to differ by *Mtb* strain and lineage^5–7^ and may be mitigated by compensatory mutations^8,9^. For example, strains belonging to the *Mtb* lineage 2 (L2) Beijing sub-lineage have been associated with a higher rate of drug resistance acquisition without a reduction in fitness^10,11^, and appear to be a major cause of MDR-TB transmission in many settings. In contrast, the globally dispersed *Mtb* lineage 4 (L4) appears to exhibit more variability in transmissibility between the distinct sub-lineages^12^.

We previously reported extensive local transmission of MDR-TB in the Republic of Moldova driven by three large clades of of highly drug resistant strains^13^. Two clades of Beijing lineage 2.2.1 strains were found mainly in the semiautonomous region of Transnistria and the third Ural lineage 4.2 clade was present throughout the country. While the Ural lineage has been reported in other former countries of the Soviet Union^14,15^, it has not previously been associated with significant local transmission^10^. This apparent widespread transmission of MDR Ural strains in Moldova is a concerning finding.

Here we compare the predicted fitness of MDR and pan-susceptible Ural and Beijing sub-lineage strains from the Republic of Moldova. Using phylodynamic approaches, we estimate the fitness of clades of MDR and non-MDR strains from both sub-lineages. We identify key genomic differences between these groups to explore putative mechanisms for transmission success of MDR-TB strains in the region. The genomic characterization of MDR-TB strains is critical for the identification of strains with epidemic potential and can aid in the surveillance of such strains throughout the region.

## Results

### Sample Data

We use data from a previously reported prospective countrywide study of *Mtb* in the Republic of Moldova^13^. These data include demographic data on all culture-positive TB cases among non-incarcerated adults in the country over the period 1 January 2018 to 31 December 2019 and whole genome sequencing of clinical isolates collected at the time of diagnosis. Of 2770 individuals diagnosed over the study period, 2236 consented to participate in the study and had a *Mtb* isolate available for sequencing. Sequence data was obtained for 2220 patients; 386 individuals were excluded due to evidence of polyclonal infections and 1834 patients were included in the final dataset.

An *in silico* approach was used to predict lineages and antimicrobial resistance profiles of all strains in the collection^16^. These predictions aligned with phenotypic testing where available (Supplementary Table 1). There were 804 *Mtb* sequences of the Beijing lineage 2.2.1 (44%), 420 sequences of the Ural lineage 4.2 (23%), and 594 sequences (32%) of other L4 strains. We focused our analyses on Beijing lineage 2.2.1 and Ural lineage 4.2 as these strains were responsible for 97% of the MDR-TB observed in Moldova over the study period.

### Clade identification

Timed phylogenetic trees were constructed separately for the Beijing lineage 2.2.1 and Ural lineage 4.2 sequences, calibrated by collection dates at the tips (Figure 1). Next, we looked to identify groups in each tree with distinct demographic or epidemiological histories to conduct phylodynamic analyses and predict clade-level expansions. We used a time threshold to characterize clades that emerged within the last 120 years on the phylogeny, censoring any clades smaller than 20 taxa. The Ural lineage 4.2 taxa were divided into three clades using this time-based approach (Figure 1A), with two of these clades containing mostly non-MDR strains (Ural_B 1% MDR and Ural_C 0% MDR), and one clade almost completely comprising MDR strains (Ural_A 95.8% MDR). This Ural_A clade also includes almost all MDR Ural strains included in the study population (252/256, 98.4%). With the time-based approach, the Beijing lineage 2.2.1 taxa were divided into multiple clades that are more heterogenous than the Ural clades in terms of the proportion of MDR strains (Figure 1B). We also used an alternative approach to define clades that detects cryptic population structure within phylogenetic trees called *TreeStructure*^17^. This model compares the observed ordering of ancestors to the expected ordering in a homogenous population. With this method, we identified population structure in the Ural lineage 4.2 tree (Supplementary figure 1A), but not in the Beijing lineage 2.2.1 tree (Supplementary figure 1B).

**Figure 1.**
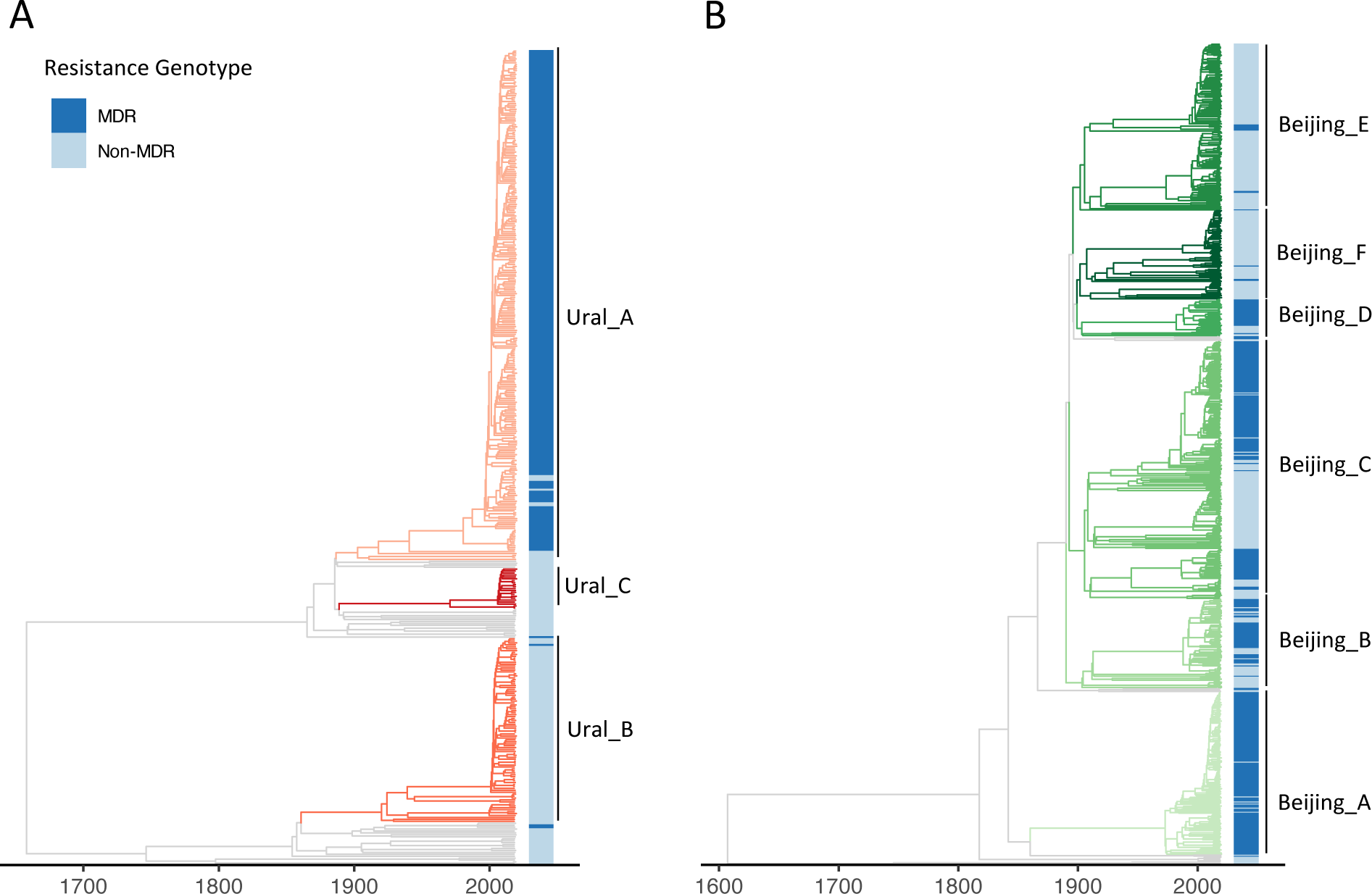
Timed phylogenetic trees of Ural (A) and Beijing (B) strains used in the study. Branches are coloured by the time-based clade designations with corresponding annotated clades names (right). MDR (dark blue) and non-MDR (light blue) strains are indicated with color.

### Markers of clade expansion

To investigate comparative rates of expansion in the Ural and Beijing time-based clades, we estimated the Local Branching Index (LBI)^18^ from a maximum likelihood phylogeny produced using the full *M. tuberculosis* dataset, including taxa from both lineages. LBI uses the length of the branches around each internal and terminal node in a phylogenetic tree to estimate local clade expansions, with large LBI values consistent with more rapid branching and clade growth. Comparing LBIs of terminal nodes, we found that the LBIs of Ural lineage 4.2 MDR taxa were higher on average than Ural lineage 4.2 non-MDR taxa and Beijing lineage 2.2.1 taxa (ANOVA p < 0.001) (Figure 2A). LBI values for Ural lineage 4.2 MDR taxa were also higher on average than LBIs for taxa belonging to other L4 strains in the collection (Supplementary figure 2). Concordantly, the mean LBI in the majority MDR clade Ural_A was 0.026, higher than clades Ural_B (mean LBI = 0.011, p < 0.001) and Ural_C (mean LBI = 0.008, p < 0.001) (Figure 2B). In contrast, the mean LBI of the MDR clade Beijing_A was 0.022, lower than the mean LBI estimate across the remaining clades (0.025, t-test p < 0.001).

**Figure 2.**
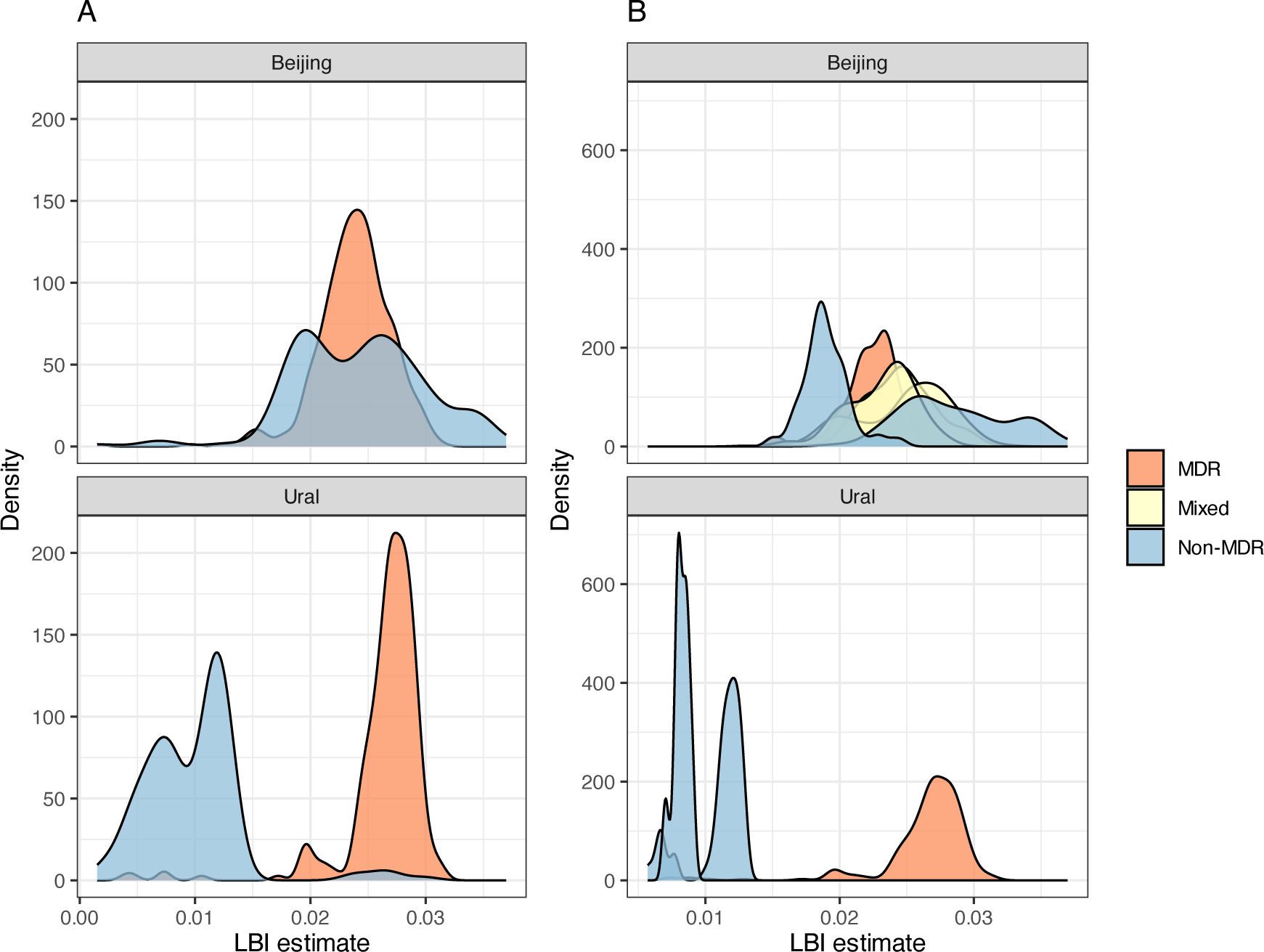
(A) Local branching index (LBI) values for all MDR (orange) and non-MDR (blue) strains used in the study for the Beijing and Ural lineages. (B) LBI values for strains in each time-based clade, colored by whether the clade is majority MDR (≥ 90% - orange), non-MDR (<10% - blue), or mixed MDR and non-MDR (yellow).

Next, we used SkyGrowth^19^ to estimate effective population sizes (N_e_) of each time-based clade separately in the timed phylogenies. This nonparametric method models the growth rate of the effective population size over discrete time intervals, which has a direct relation to the effective reproduction number (R_e_). We predicted the time dependent N_e_ between 1990 and the beginning of sampling in 2018 for each clade (Figure 3). We found that the N_e_ of the majority MDR clade, Ural_A, increased 3.0 fold (95% CrI: 1.30, 5.76) over the period 2010-2018, indicating the recent rapid expansion of this clade. The Ural_A increase in N_e_ was markedly higher than that of the majority non-MDR Ural clades (Ural_B 0.26 [-0.45, 2.1] and Ural_C -0.29 [-0.84, 2.09]), as well as the majority MDR clade, Beijing_A, (1.18 [0.16, 3.12]), over the same approximate time period.

**Figure 3.**
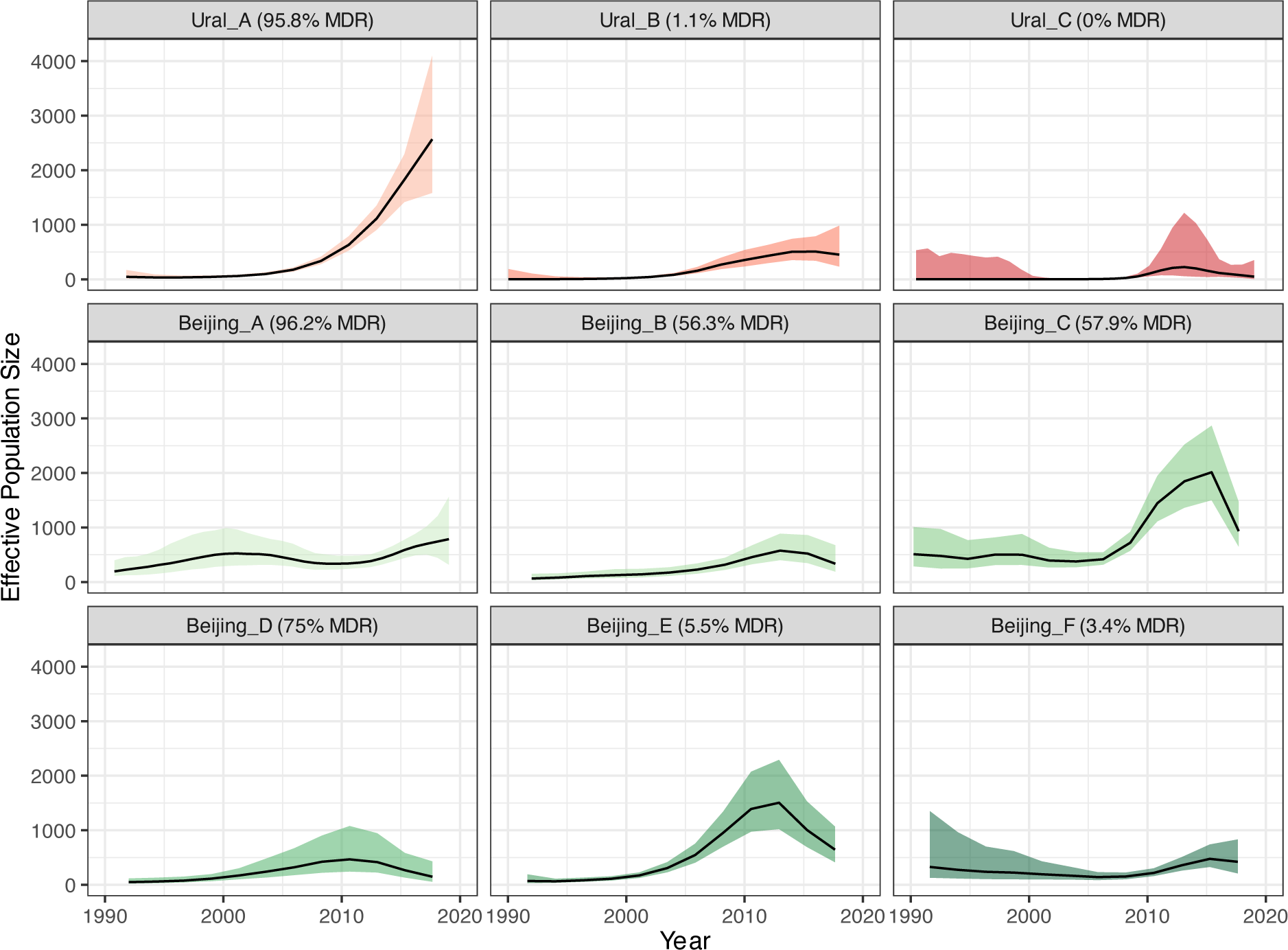
Effective population size estimates for each time-based clade, inferred using *SkyGrowth*. Ural lineage clades are in red and Beijing lineage clades in green, with the percentage of MDR strains in each clade shown in the panel header.

### The effective reproduction number of Ural strains

We estimated the effective reproduction number (R_e_) of MDR and Non-MDR Ural strains using a multi-type birth death (MTBD) model implemented in the Bayesian phylodynamic inference software BEAST2^20^. R_e_ is the expected number of secondary infections caused by a single infectious individual; a higher R_e_ suggests a pathogen is spreading more rapidly. Under a birth-death model, new infections are ‘born’ when a new host is infected, ‘die’ when the host recovers, and are sampled with some probability (zero before the beginning of the study period). Infections are stratified into several ‘types’, and the effective reproduction number of each type is the birth rate divided by the death rate. We found that the predicted R_e_ of the MDR Ural lineage 4.2 strains was R_e_ = 2.5 (95% HPD: 1.59, 3.80). This was substantially higher than the non-MDR Ural 4.2 strains, with an estimated R_e_ = 1.06 (1.02, 1.16); across model iterations, the R_e_ of MDR Ural strains was 2.34 (1.55, 3.40) times higher than non-MDR Ural strains.

**Fig 4:**
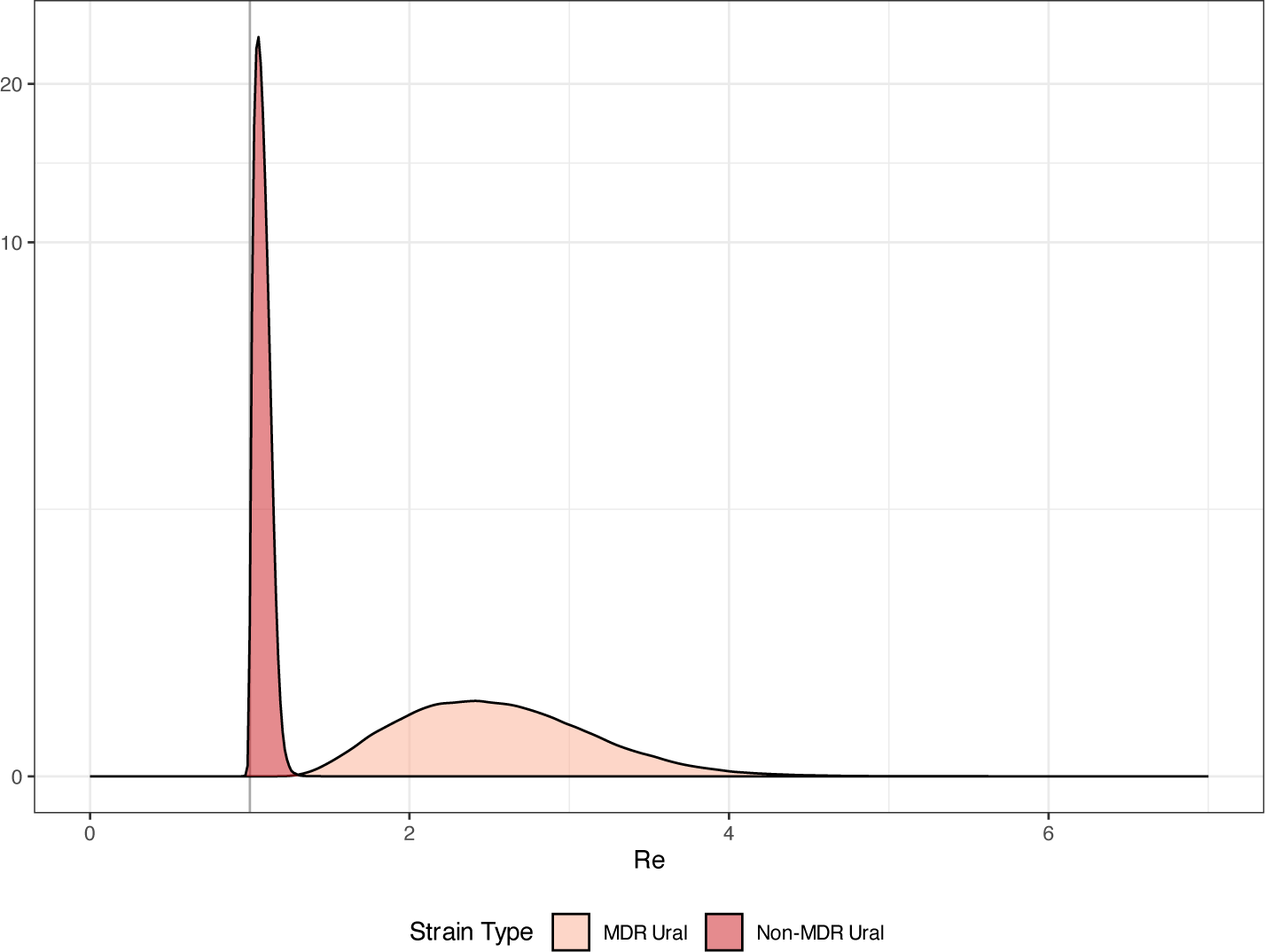
The effective reproduction number (R_e_) of MDR (light red) and non-MDR (dark red) Ural strains inferred using a multi-type birth death model in *BEAST2*.

### Genomic associations with Ural MDR-TB in Moldova

We characterized mutational differences between the MDR Ural isolates and the non-MDR Ural lineage 4.2 and MDR Beijing strains. We identified 70 SNPs and 5 short insertions or deletions (indels) with an allele frequency of ≥ 90% in the Ural MDR strains that occurred at a frequency of < 10% in the non-MDR Ural or MDR Beijing strains (Supplementary table 2). Of the 70 SNPs, 37 were non-synonymous mutations in coding regions, 29 were synonymous coding SNPs and four were in intergenic regions.

There was evidence of multiple non-synonymous SNPs in genes that have been previously linked to antimicrobial resistance. The *rpoB* S450L mutation, which was found in high proportions in both MDR Ural and Beijing isolates and in low frequencies in non-MDR strains, has been previously well-characterized as conferring rifampin resistance^21^. We also found a high proportion of MDR Ural strains harbored a mutation in *rpsL*, K88R, that has been implicated in streptomycin resistance^22^, though with an associated fitness cost^23^. Additionally, a mutation in *ethA*, H281P, was identified in a high proportion of the MDR Ural strains and is almost absent in the rest of the population. This SNP has been observed previously in MDR strains from Moldova^24^, and mutations in this gene have been linked to resistance to the second-line TB antimicrobial, ethionamide^25^. Interestingly, we indentified a non-synonymous SNP that was present in 98% of our Ural MDR strains in *murA*, a key gene in the synthesis of an essential componenet of the *Mtb* cell wall, peptidoglycan (PG)^26^. A recent study described a previously unknown mechanistic link between PG synthesis and mutations in *rpoB* ^27^. The location of the *murA* R110T mutation, near a known active site residuce, raises the potential that this may be a compensatory mutation associated with rifampin resistance.

A synonymous SNP in *esxO* at position 2625924 (codon 83), which was fixed in the Beijing lineage, was found to be present in 94% of the MDR Ural strains and only 4% of the Ural susceptible strains. We also identified a synonymous SNP in *esxP* at codon 10 almost exclusively in the MDR Ural strains. The ESX gene family have been shown to play a role in host-pathogen interaction^28^ and we found previous evidence of a mutation in *esxW* in the Beijing strains circulating in Moldova^13^. Synonymous SNPs in the *esxO* and *esxP* have been previously identified as potentially under selection in MDR-TB strains elsewhere^29^. Additional non-synonymous SNPs were identified in genes that may confer a growth or survival advantage, including Rv0355c (*PPE8*) that has been associated with adaptation to host defense mechanisms^30^, *mmaA4* previously implicated in drug resistance through inhibition of the mycolic acid biosynthetic pathway^31^, and Rv1835c that appears to confer a growth advantage in *Mtb* when disrupted based on transposon mutagenesis^32^. Furthermore, we found five indels at a high frequency in MDR Ural strains that were not present in other isolates, including 2bp frameshift insertion in the *ndhA* gene, though this has been previously reported to be a non-essential gene for *Mtb* survival^33^.

Finally, we performed a genome-wide association study (GWAS) to identify SNPs associated with multiple drug resistance in both the Ural and Beijing sublineages (Supplementary table 3). In the Ural strains, we identified only the *rpoB* S450L mutation as significantly associated with MDR strains. We found the same mutation in the Beijing strains, along with the *katG* S315R mutation, which confers isoniazid resistance^34^, and the *rpsL* K43R mutation, which confers streptomycin resistance^22^.

### Evidence of highly transmissible Ural MDR-TB strains outside of Moldova

The Ural lineage 4.2 has been identified in other countries of the former Soviet Union^10,35^ and in light of this, we compared our strains to publicly available *Mtb* sequences from Georgia^10^. Although the fraction of MDR strains among Ural strains isolated in Georgia was far lower than in Moldova (∼6% MDR in Georgia compared to ∼61% MDR in Moldova), there was evidence that nine of the 27 MDR Ural isolates from Georgia had similar mutational profiles to our MDR Ural strains from Moldova (Supplementary table 2). Of the 70 MDR Ural strain-specific SNPs identified above in Moldovan isolates, 59 (84%) of these SNPs were found in approximately one third of MDR Ural strains from Georgia (9 strains), and four of these strains also harbored a further 7 SNPs (94% of the total SNPs). Two of the known AMR associated SNPs in *rpoB* and *rpsL* were found in higher frequencies in the Georgian MDR Ural strains (78% and 52% respectively) and only two SNPs were not present at all in the MDR Ural isolates from Georgia. All the 70 MDR Ural strain-specific SNPs identified in Moldovan samples were found at a low frequency in pan-susceptible Ural strains from Georgia (< 10%).

### Sensitivity Analyses

We conducted several additional analyses to determine whether these results were sensitive to modeling assumptions. To account for the evidence that the average mutation rate in *Mtb* L4 is believed to be slower than in Beijing strains^4^, we reconstructed an additional timed phylogenetic tree for the Ural sub-lineage 4.2 with a fixed mutation rate of 0.3 SNPs/genome/year and recalculated effective population sizes. We found that tree could be subdivided into time-based clades that were similar in size and composition to the those identified in the main analysis, and that the majority MDR Ural clade was still predicted to have a faster growth rate than the non-MDR Ural clades and the Beijing clades in the main analysis (Supplementary figure 3). We also randomly resampled the Ural sub-lineage 4.2 sequences and re-ran the MTBD model. We found that R_e_ estimates were consistent across the main analysis and the re-sampled run (Supplementary figure 4). Finally, we looked at the demographic composition of the timed clades used in this study to determine whether these factors may contribute to the observed differences in transmission. We found that clades had similar distributions of age, sex, and homelessness (Supplementary table 4). The Bejing_A clade had a higher proportion of formerly incarcerated individuals (24.5%) than the overall average (11.4%), though all other clades did not vary greatly in their fraction of formerly incarcerated individuals (Supplementary table 4).

## Discussion

We investigated the reproductive fitness of the *Mtb* Ural 4.2 and Beijing 2.2.1 sub-lineage strains responsible for most MDR-TB circulating in the Republic of Moldova. We found that the fitness of MDR Ural strains is high relative to pan-susceptible strains of the same lineage, and even higher than strains of the Beijing sub-lineage commonly associated with successful transmission. We also found strong evidence that there has been a recent, rapid expansion of a large MDR clade of Ural lineage 4.2 strains in Moldova in the past 10-15 years. Given the high burden of MDR-TB in Eastern Europe, these results suggest that MDR-TB of the Ural 4.2 sub-lineage should be closely monitored in Moldova and the surrounding region to enhance TB control efforts and prevent widespread transmission.

Our findings are consistent with our earlier work in the Republic of Moldova, which highlighted the presence of highly drug resistant Ural lineage 4.2 strains that appeared to be readily transmitting within the population^13,36–38^. However, the R_e_ estimates for Ural 4.2 strains reported here contrast with a recently published study that found that there was a reduced transmission fitness associated with multi-drug resistance in L4 strains in another former Soviet Union country^10^. This Georgian study, conducted on strains collected two years prior to our work in Moldova, identified only 27 MDR Ural 4.2 strains, and only 14% of the MDR-TB in the country was in L4 strains. Notably, we found that a small number of the Georgian MDR Ural strains had similar mutational profiles to the Moldovan MDR Ural strains identified in our study. Given the evidence of the recent expansion of MDR Ural strains in Moldova in the past decade, it is possible that the MDR Ural sub-lineage may now be circulating more widely in Georgia and other countries of the former Soviet Union.

We identified several genes associated with anti-tuberculosis drug resistance in the MDR Ural strains that were not present in high proportions in MDR Beijing strains, including genes conferring resistance to second-line drugs streptomycin and ethionamide. In addition, we identified several mutations that are associated with improved bacterial survival, including in ESX family of genes which are involved in host-pathogen interactions^39^. While these findings provide potential mechanisms by which the relative fitness of the MDR Ural strains is maintained or increased, we only looked for the presence of single variants that were at a high frequency in our MDR strains. A more comprehensive investigation could provide further insights, including the specific fitness effects conferred by individual drug-resistant mutations and putative compensatory mutations, along with an analysis of epistatic interactions between multiple variants present concurrently in MDR strains and the functional modelling of protein changes.

A limitation of this study was that our dataset included *Mtb* isolates collected over a two-year period, making it difficult to observe longer term trends in strain evolution. For this reason, our data are not sufficient to estimate a mutation rate; we used a fixed mutation rate to estimate time-resolved phylogenies. This choice may impact estimates of the effective population size. We reconstructed the timed phylogeny with the Ural strains using a lower fixed mutation rate and found the same pattern of rapid expansion in the majority MDR Ural clade. Furthermore, the reported clade expansion estimates were inferred using clades that were defined by applying a cut-off to the timed phylogeny of 120 years before the study period, removing any tips with a coalescent time to the clade beyond this threshold. As such, these results may be biased by excluding MDR strains with low transmission fitness and by the specific choice of threshold. Even so, the final clades include the majority of Ural and Beijing strains from our dataset and the overall finding, that MDR Ural strains appear to have a high transmission fitness than susceptible Ural strains, is supported by the LBI estimates that included all strains in the collection.

In conclusion, we found that MDR-TB Ural lineage 4.2 strains have significantly higher effective reproductive fitness than non-MDR Ural strains circulating in the Republic of Moldova. There is also evidence of a recent, rapid expansion of a large clade of MDR Ural strains, in contrast to non-MDR Ural and both MDR and non-MDR Beijing clades that do not show the same concerning trajectory. Given that we find that approximately 18% of Ural strains in our study were resistant to fluroquinolones, there is also the risk that the MDR-TB Ural lineage 4.2 strain develops further drug resistance and could evolve to become extensively drug resistant^40,41^. Tracking the evolution and transmission of MDR-TB Ural strains in countries of the former Soviet Union is vital to reduce the burden of MDR-TB in these regions and to implement effective TB control strategies.

## Online Methods

### Study Population and Sequence Analysis

Between January 2018 and December 2019, TB culture-positive individuals in the Republic of Moldova were asked to consent for the use of routinely collected demographic and epidemiological data, and for whole genome sequencing of bacterial isolates. Ethical approval was obtained for this study from the Ethics Committee of Research of the Phthisiopneumology Institute in Moldova and the Yale University Human Investigation Committee (Number 2000023071). Next generation sequencing was carried out using the Illumina MiSeq platform, with the resulting raw reads mapped to the H37Rv reference strain with *BWA* ‘mem’^42^ and single nucleotide polymorphisms (SNPs) called using the GATK software package^43^. We identified polyclonal infections using *MixInfect*^44^ and excluded these samples from further analysis. Lineage typing and *in silico* drug resistance predictions were carried out with *TB-Profiler* v2.8.14^16^. Full details on the study enrollment, specimen and individual metadata collection, and whole genome sequencing have been described previously^13^.

### Phylogenetic tree construction and clade identification

A maximum likelihood (ML) phylogeny comprising all isolates was produced from a multiple sequence alignment of concatenated SNPs using *RAxML-NG*^45^ with the ‘GTRGAMMA’ substitution model. We next constructed time-calibrated phylogenies separately for Beijing lineage 2.2 and Ural lineage 4.2 strains using the R package *BactDating*^46^ and the well-resolved ML tree as input. Timed phylogenies were built using the ‘strictgamma’ clock model, a root finding algorithm,and a fixed mutation rate 0.5 SNPs/genome/year, which corresponds to previously reported estimates for the *M. tuberculosis* mutation rate^4^. We also conducted a sensitivity analysis by repeating the analysis on the Ural lineage 4.2 strain using a fixed mutation rate of 0.3 SNPs/genome/year to reflect the potentially slower rate reported for Mtb lineage 4. We ran *BactDating* for 1×10^7^ Markov Chain Monte Carlo (MCMC) iterations, thinning by a factor of 10,000, without a coalescent prior. We assigned taxa to clades with a shared evolutionary history using two different phylogenetic methods. First, we used a time-based method to identify clades with 20 or more tips that emerged within 120 years start of the study period. Second, we assigned taxa to clades based on evidence of an underlying population structure in the phylogeny using the R package *treestructure*^17^.

### Phylodynamic Analyses

We estimated the Local Branching Index (LBI) of each tip in the ML phylogeny using the R package *TreeImbalance*^47^. LBI is a quantitative method to estimate fitness from the shape of phylogenetic tree. LBI estimates are based on the total branch length surrounding each internal and terminal node in a phylogeny, discounting exponentially with increasing distance from the node. Higher values of LBI are associated with rapid expansion and increased fitness. The scale of the exponential discounting, τ, is a function of the relevant neighborhood size around a node. Neher et al. report that τ should be the average pairwise distance within the tree scaled by a factor of 0.0625^18^. We calculate the LBI for each internal and terminal node based on the ML tree and the scaling factor τ and summarize results for each terminal node by drug resistant genotype and by clade.

Next, we estimated effective population sizes (N_e_) using the R package *skygrowth*^19^, a nonparametric autoregressive model to estimate effective population sizes through time. We fitted the model to each time clade separately with a Bayesian MCMC approach and ran the model for 100,000 iterations, allowing estimation of N_e_ over 50 timepoints prior to the beginning of sampling and a precision factor of 1 standard deviation.

We directly inferred the effective reproduction numbers (R_e_) for MDR and non-MDR Ural lineage 4.2 strains using a multi-type birth death (MTBD) model ^48^. We sampled 200 taxa from the Ural lineage 4.2, maintaining the same proportion of MDR and non-MDR taxa as in the full dataset. The *bdmm* package^49^ in BEAST2^20^ version 2.7.4 was used to run the MTBD model using the multisequence alignment of concatenated SNPs and collection dates, accounting for invariant sites. We ran the model for 100 million MCMC iterations or until all parameters had an effective sample size greater than 200.

### Genomic Analysis

We identified mutational differences between the MDR and non-MDR strains Ural strains and the MDR Ural and MDR Beijing strains by comparing the allele frequency of all SNPs and short insertions and deletions in these groups. We examined genomic differences between the MDR Ural strains from Moldova and known antimicrobial resistance mutations were annotated using in-house lists based on associated variants described previously^16,50^ (Supplemental table S5). Finally, we conducted a GWAS to find SNPs associated with MDR in both the Ural and Beijing strains using *treeWAS*^51^.

## Data Availability

All data produced in the present study are available upon reasonable request to the authors

---

**Supplementary figure 1.**
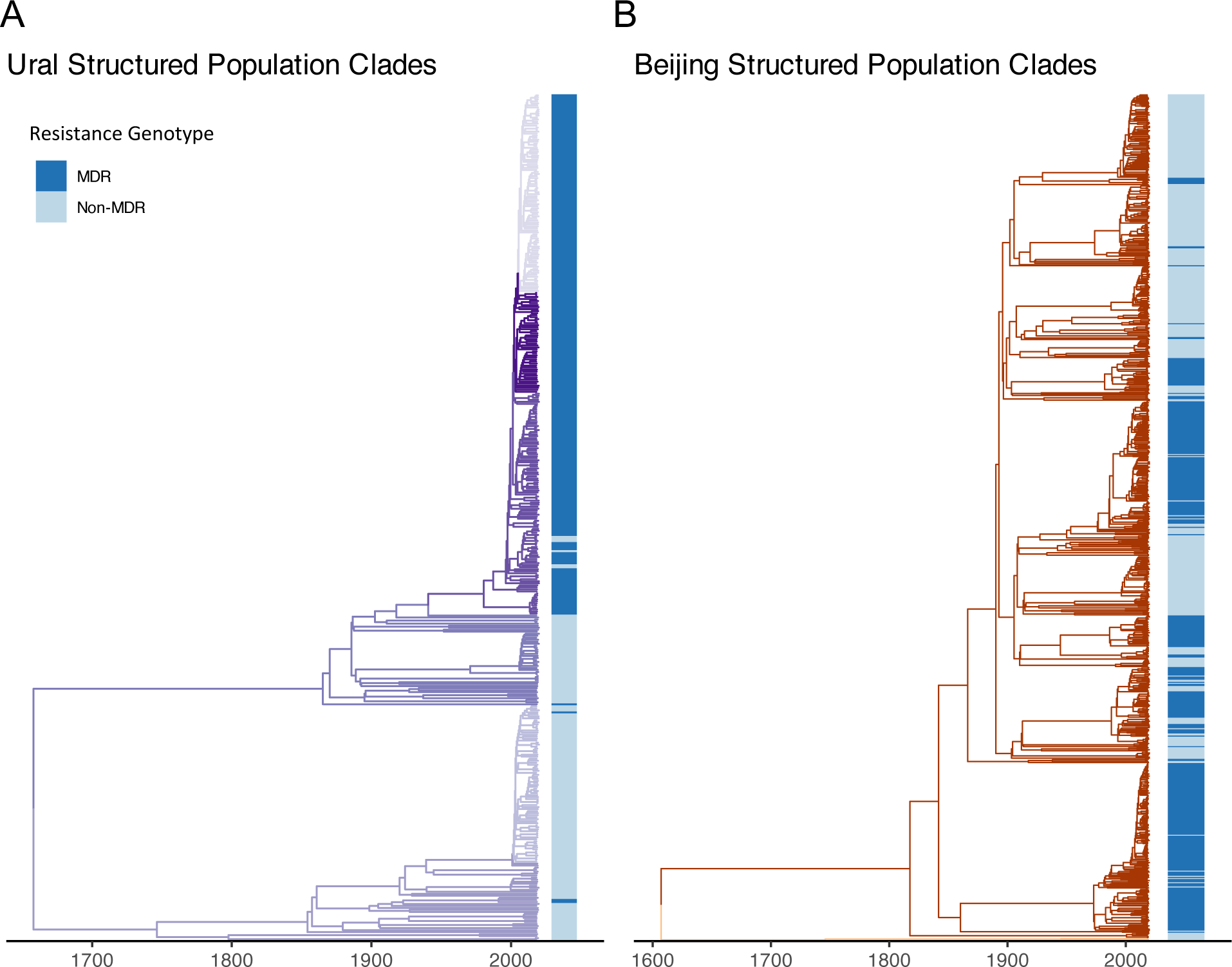
Timed phylogenetic trees of Ural (A) and Beijing (B) strains used in the study. Branches are coloured by structured population clade designations estimated with *TreeStructure*, and the position of MDR (dark blue) and non-MDR (light blue) strains shown in the coloured bar.

**Supplementary figure 2.**
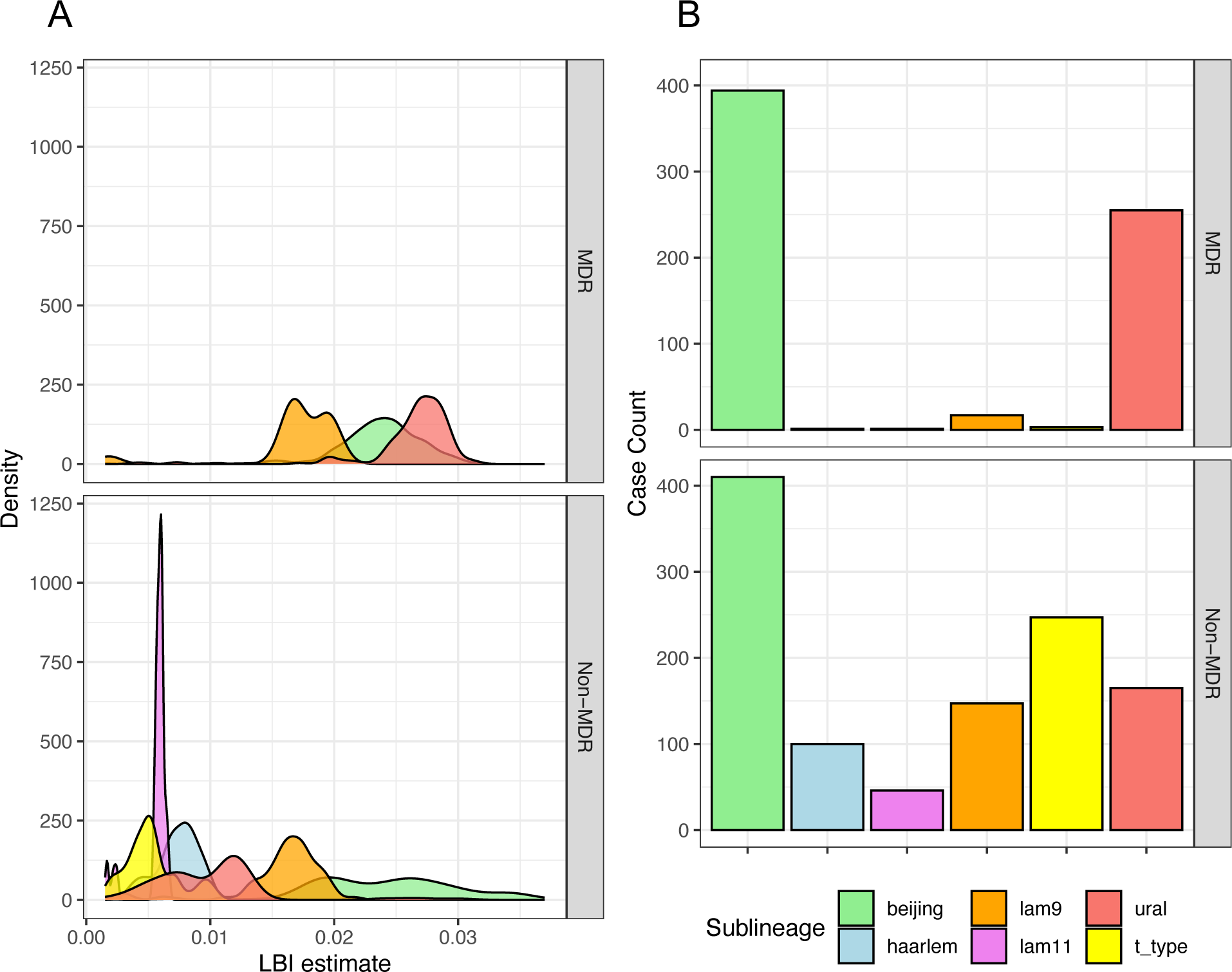
(A) Local branching index (LBI) estimates for all lineage 2 and lineage 4 sub-lineages present in Moldova during the study period, separated by MDR and non-MDR strain status. (B) Case counts of sequences collected during the study period of each lineage2 and lineage 4 sub-lineage, separated by MDR and non-MDR strain status.

**Supplementary figure 3.**
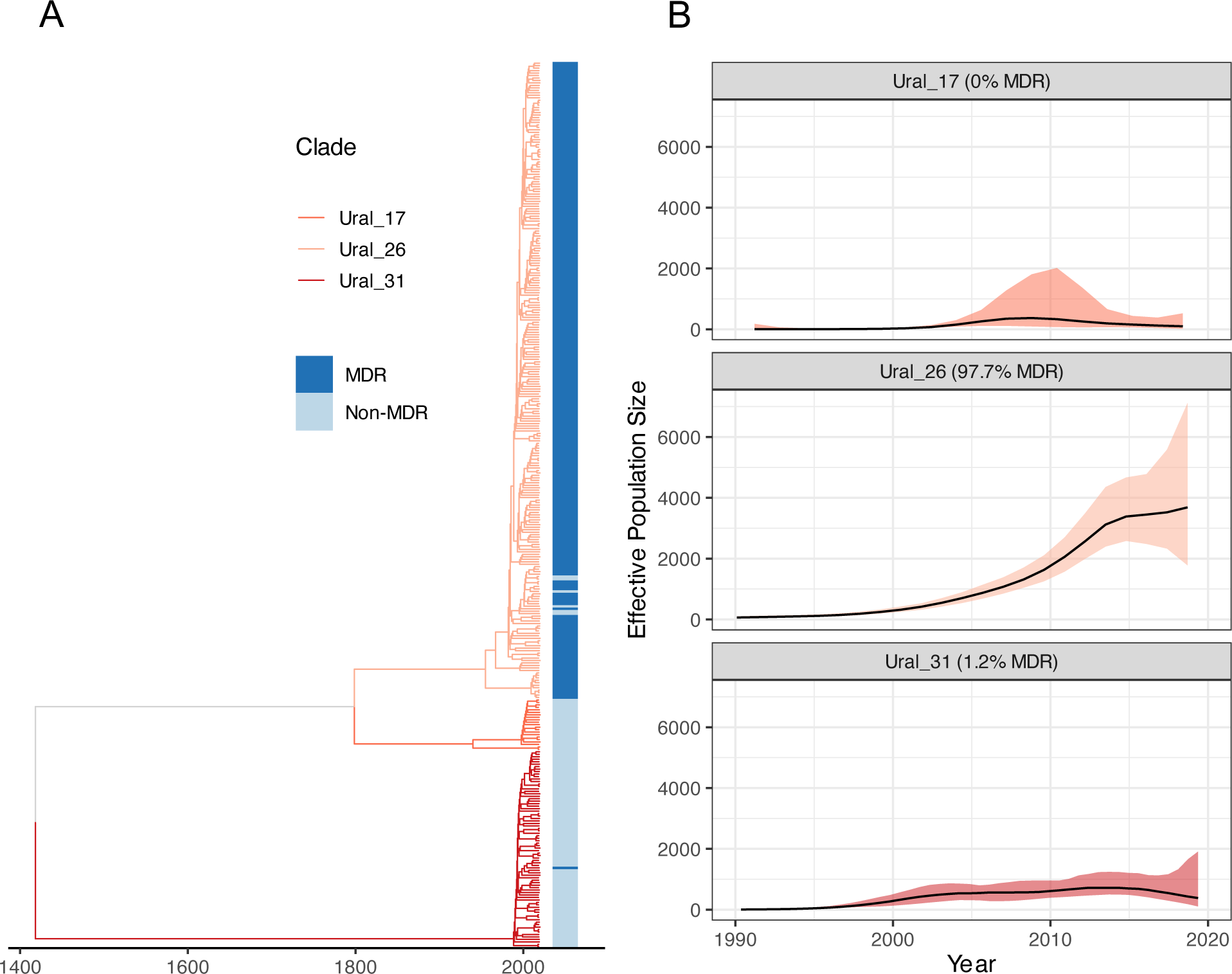
(A) Timed phylogenetic tree of Ural strains reconstructed using a lower mutation rate of 0.3 SNPs/genome/year, compared to 0.5 SNPs/genome/year in the main analysis, with MDR status and new time-based clade designation shown. (B) Effective population size estimates for each new time-based clade, inferred using *SkyGrowth*, showing the rapid growth of a majority MDR Ural clade (Ural_26), with corresponds to the Ural_A clade in the main analysis.

**Supplementary figure 4.**
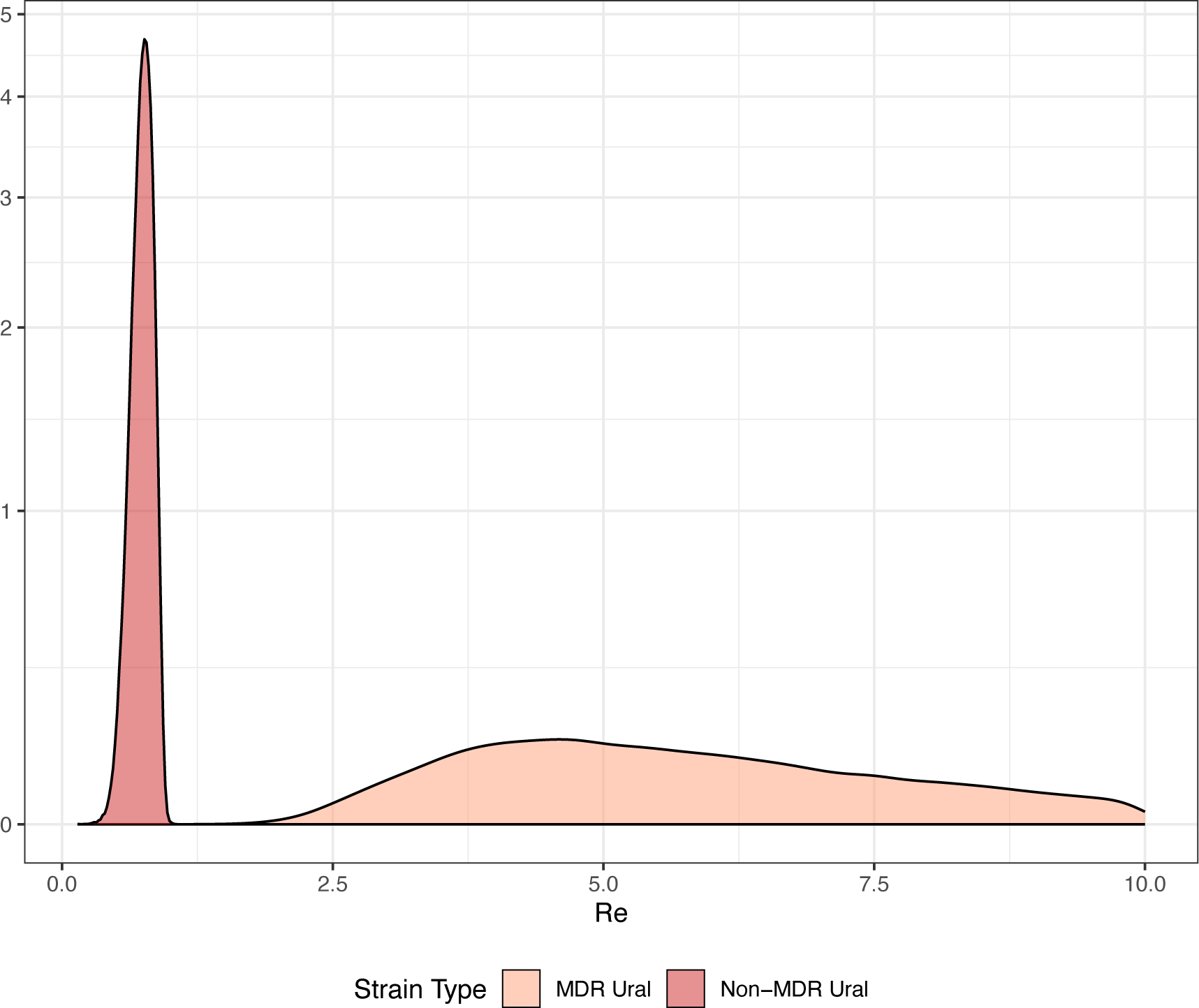
The effective reproduction number (R_e_) of MDR (light red) and non-MDR (dark red) randomly down-sampled Ural strains inferred using a multi-type birth death model in *BEAST2*.

